# Rapid transition to distance learning due to COVID-19: Perceptions of postgraduate dental learners and instructors

**DOI:** 10.1101/2020.09.19.20197830

**Authors:** Fatemeh Amir Rad, Farah Otaki, Zaid Baqain, Nabil Zary, Manal Al-Halabi

## Abstract

The outbreak of COVID-19 necessitated abrupt transition from on campus, face-to- face sessions to online, distance learning in higher education institutions. The purpose of this study was to investigate the perceptions of postgraduate dental learners and instructors about the transition to distance learning, including the changes to the learning and teaching and its efficaciousness.

A mixed-methods approach to research was utilized. All the instructors and postgraduate learners were invited to participate in the online survey. Quantitative data was analyzed using descriptive and inferential analyses on SPSS for Windows version 25.0, and for the responses to the open-ended questions, multi-staged Thematic Analysis was utilized.

Both groups of stakeholders: learners and instructors, were quite satisfied with the rapid transition to distance learning due to COVID-19. Instructors were significantly more satisfied than the learners. The stakeholders adapted well to the change. The perception of the stakeholders regarding the case-based scenarios significantly influenced their level of satisfaction. As perceived by the stakeholders, the transition to distance learning entailed advantages and challenges. Going through the experience equipped the stakeholders with lessons learned and enabled them to develop informed opinions of how best to sustain learning and teaching irrespective of how matters unfold in relation to the pandemic.

In conclusion, the worldwide dental education community faced unprecedented challenges due to the onset of COVID-19. Yet, in the grand scheme of things, it is important for decision-makers not to miss-out on the worthwhile opportunities, inherent in the experience, to reinforce curriculums, and maximize the learning and teaching.

## Introduction

It took the coronavirus disease 2019 (COVID-19) two months to traverse national borders, across multiple continents. On the 11^th^ of March 2020, the World Health Organization (WHO) announced that the COVID-19 epidemic transformed into a pandemic (1). The outbreak of COVID-19 led to a rapidly evolving situation which impacted the education system worldwide (2). Continuing the delivery of education through alternative learning and teaching channels abruptly became a top priority for institutions aiming to keep the impact of the crisis on education to a minimum. Following the endorsement of national social distancing directives, education institutions, across many countries, had no option but to resort to distance learning environments and other e-learning resources (3). Implementation of distance learning, in the United Arab Emirates (UAE), for all higher education institutions’ became a requirement starting from the 22^nd^ of March, 2020 until the end of June, 2020, where almost three quarters of the second half of 2019-2020 academic year was conducted online (4).

Rapid transition from on campus, face-to-face classes to online, distance learning sessions took place. Many educators had never delivered sessions via online environments, which required them to acquire an extensive set of skills over a short period of time. They also needed to adapt the content and structure of their offerings, and to select the most suitable methods to engage their learners in the virtual environment. The impact of these restrictions were exacerbated, in health professions education, due to the suspension of all elective treatments where the associated experiential education constitutes the core of the learning and teaching (5). There are pedagogical approaches for planning of distance learning courses, which require special techniques of course design, instructional design, and methods of communication (6, 7). Yet, given the urgency of the COVID-19 situation, institutions did not get the space to plan for, and undergo the proper, systematic way of transitioning, which usually involves substantial amount of capacity building (i.e., offering learning and development opportunities for involved stakeholders) and of change management (i.e., striving to minimize the inevitable resistance and its counterproductive consequences to any institutional changes). In addition, the learners who were accustomed to face-to-face interactions, had to rapidly adapt to distance learning and to the online environment. Moreover, the challenges around rapidly transitioning to distance learning were exacerbated by the multiple changes and restrictions accompanying COVID-19, and the resulting psychosocial stressors that the learners and the educators has been facing (8, 9). Educators and learners needed to increase networking, foster the humanity in their connections, and enhance the effectiveness of their communication before, during, and after their online engagements (10). This experience influenced the way they create meaning and reflect upon the learning and teaching (11).

The novelty of the situation and how the involved parties have been adapting to this situation constitute worthy opportunities for investigation given that most research studies, to date, have been conducted in “typical” (relatively more stable) educational environments. It is important to examine and reflect on this experience to better prepare for the potentiality of reoccurrence and the experiencing of other similar emergency situations. Moreover, the lessons learned from this sudden transition of adults’ education hold the potential of positively transforming post-pandemic learning and teaching, especially in programs, that rely heavily on clinical training, since they were impacted the most since the onset of this pandemic. This is particularly relevant to post-graduate dental education since the learning and teaching is heavily reliant on clinical specialty training.

The purpose of this study was therefore to investigate the perceptions of postgraduate dental learners and instructors in relation to the transition to distance learning, including the changes to the learning and teaching and its efficaciousness, and how these stakeholders modified their learning or teaching to adapt to this abrupt change.

Accordingly, in this study, we strive to address the following research question: how was the rapid transition to distance learning due to COVID-19 perceived by postgraduate dental learners and instructors, and how do those perceptions relate to one another?

## Materials and methods

### Context of the study

This study was undertaken at Hamdan Bin Mohammed College of Dental Medicine (HBMCDM) at Mohammed Bin Rashid University of Medicine and Health Sciences (MBRU), Dubai, UAE. HBMCDM is a new postgraduate dental school, launched in Academic Year 2013-2014, that offers three-years full-time specialty dental postgraduate programs in endodontics, orthodontics, pediatric dentistry, periodontics, and prosthodontics.

### Description of the transition to distance learning

In an effort to curtail the spread of the COVID-19 pandemic and in response to the directive of the Ministry of Education (Decree 229)(12), HBMCDM along with all other educational institutions in the UAE switched to complete distance learning as of the 22^nd^ of March, 2020 until the end of the academic year (the 9^th^ of July, 2020 in the case of HBMCDM- the college under investigation in this study). All didactic educational activities were continued as scheduled. At the time of transition, the first-year learners were preparing their research protocols, and the third-year learners were mainly engaged in the preparation of the graduate dissertation. Both of those groups of learners were able to pursue their research work. As for some of the second-year learners, had to stop the empirical (i.e., data collection) part of their studies given the implemented COVID-19 directives. The learners’ discussions and advising sessions related to the different stages of the scientific research method of the dissertations that continued (ranging from research conceptualization to determining of research methodology, and data collection tools and analyses, all the way to preparing of the manuscript) were all conducted via Microsoft Teams.

For the clinical training, it was decided that the learners will have to compensate for the missed clinical activities to ensure the attainment of the required clinical competencies at a later stage. To make-up for the generated gap, a compensation program in the following academic year was introduced. In the meantime, the college focused on virtual case-based discussion sessions and involved learners in Teleconsultations under the supervision of faculty members. In addition, some learners were deployed to off-training sites to assist authorities with COVID-19 contact tracing and sampling.

## Intervention

### Platforms

The digital platforms utilized for delivery of distance learning consisted mainly of two platforms. HBMCDM had an existing Learning Management System (LMS) that has been in function since 2014. The LMS is used by course instructors to post course information and content in addition to conducting assessments and posting grades. It is also used for interactive discussions within specific courses. The use of LMS was maintained throughout the distance learning. The heavy reliance on Microsoft Teams was novel. It constituted the main platform which provided a medium for real-time class presentations by both learners and instructors, and for research dissertation- related interactions and clinical CBD. In addition, some instructors pre-recorded their lectures with the support of the educational technologists at MBRU to ensure the learners can access the content at their convenience.

### Instructors’ professional learning and development

All instructors were required to undergo learning and development sessions, concerning the transition to distance learning, delivered by MBRU Faculty Development and Information Technology (IT) support teams. These sessions were conducted via Microsoft Teams. The IT support team assigned technical support personnel to each instructor. In addition, the Faculty Development team provided one-to-one consulting to support the instructors in learning design, where instructions were adapted to the respective instructors’ learning and teaching specifications. The instructors were advised to shorten the length of the teaching sessions, not to exceed one hour each. Instructors were also advised to provide reading material prior to the teaching sessions to enable the suggested shortening of the session.

### The teaching sessions

The length of time allocated to different classes remained unchanged. As for the scheduled timing of the lectures, the original schedule for the classes assigned at the beginning of the second semester of Academic Year 2019-2020 was used. Class attendance was registered by the course instructor on MBRU Self-Service portal.

Additional two-hour Case-Based Discussions (CBD) were added to the original two- hour CBD to have in total four-hours of CBD across two sessions per week. These sessions were meant to engage the graduate learners in certain clinical skills including diagnosis, decision making, and treatment planning through encouraging critical thinking and providing constructive multi-stream dialogue among and in between the learners and instructors.

### Changes in assessments

Major changes in assessment methods were implemented to accommodate for the absence of live proctoring. Instructors were encouraged to consider feasible alternative tracks that emphasize equity and hold the learners accountable to academic integrity. The instructors collectively needed to identify alternative means of conducting summative assessment to ensure the attainment of intended course learning outcomes and readiness of the learners to progress.

The weightage of the summative assessments for all courses as determined at the beginning of the semester was upheld. Emphasis was placed on maximizing formative Multiple-Choice Question (MCQ) type quizzes to ensure that the individual lectures’ learning outcomes were met. Instructors were encouraged to conduct assessments using clinical scenarios to test the learners’ diagnosis, clinical judgment, and problem-solving skills as well as treatment planning competencies especially in complex multidisciplinary cases. As for the oral clinical exams, which are based on unseen case-scenarios, they were conducted using Microsoft Teams.

The LMS system, through which the exams were conducted, deployed a lockdown browser requirement which prevented the learners from opening any other application on their devices while taking the exam. In addition, activation of the webcam, in the learners’ devices, was required. The system was enabled to detect any abnormal activity by the exam-taker, and flag it to be checked by the course instructor. All learners and instructors received training on how to take the exam using the lockdown browser and the webcam. In addition, detailed written instructions were sent to the learners prior to each summative exam.

### Research design

A mixed-methods study design was adopted to systematically develop an understanding of the stakeholders’ perceptions regarding the rapid transition to distance learning. This study is characterized by a single phase, where existing qualitative and quantitative data was concurrently collected and analyzed. The triangulation of data, in terms of sources (i.e., learners and instructors) and types (i.e., qualitative and quantitative), is meant to raise the validity of the generated findings.

Ethical approval for the study was granted by the MBRU, Institutional Review Board (Reference # MBRU-IRB-2020-032).

### Data collection

The data was collected using a survey that was designed specifically for the purpose of this study. It aimed at assessing the perception of learners and instructors regarding the rapid transition to distance learning due to COVID-19 and its effect on the learning and teaching at the college.

The survey was developed by three researchers (FAR, FO, and MAH), who started with looking into how the various universities across the world are planning on evaluating their distance learning performance since the onset of COVID-19. Among the universities which had their surveys readily available were the following: University of Minnesota, University of Pittsburgh, University of Saskatchewan, and Rutgers University (13). These resources were retrieved from an online community of Institutional Research professionals where an asynchronous forum discussion around “Impact of COVID-19 on evaluations in Higher Education Institutions” has been taking place. These surveys were thoroughly reflected upon, and in turn extracted segments from all got contextualized and in turn adapted for this study. The survey was composed of three segments. The first segment is a Likert scale of five points (1: Strongly Disagree, 2: Disagree, 3: Neutral, 4: Agree, and 5: Strongly Agree) across 8 variables, as per Table 1. Out of those 8 variables, 3 were replicated, as iss, for both learners and instructors, 3 others were replaced for the instructors with an alternative which is supposed to reflect the other side of the same coin, and 2 were unique to the learners.

**Table 1.**
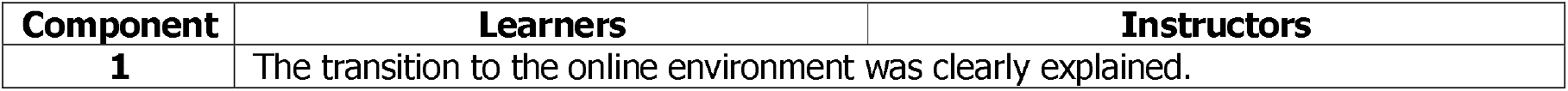
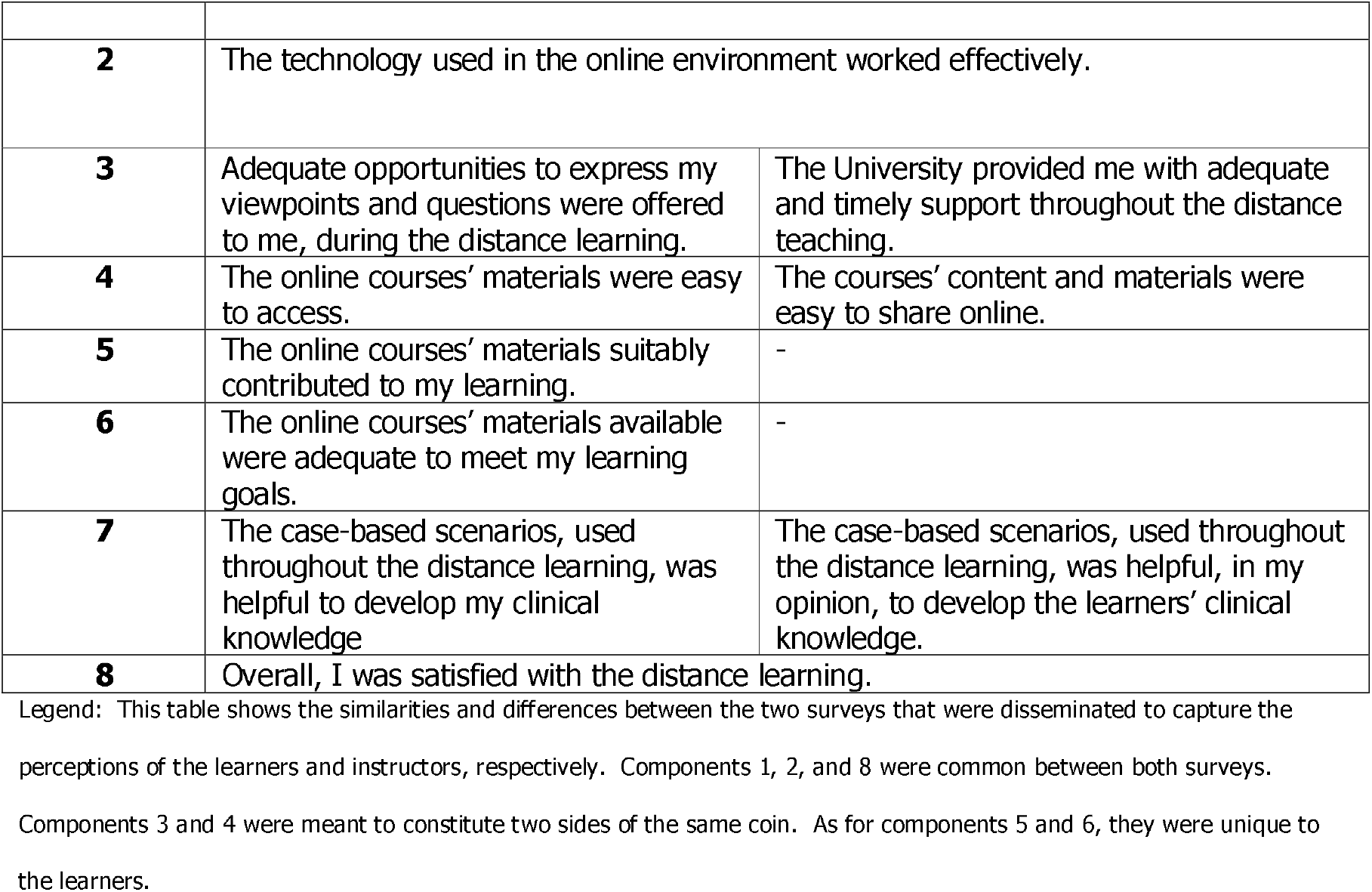
Description of the first section of the survey that captured the perceptions of the learners and instructors

| Component | Learners | Instructors |
| --- | --- | --- |
| 1 | The transition to the online environment was clearly explained. |  |
| **2** | The technology used in the online environment worked effectively. |  |
| **3** | Adequate opportunities to express my viewpoints and questions were offered to me, during the distance learning. | The University provided me with adequate and timely support throughout the distance teaching. |
| **4** | The online courses' materials were easy to access. | The courses' content and materials were easy to share online. |
| **5** | The online courses' materials suitably contributed to my learning. | - |
| **6** | The online courses' materials available were adequate to meet my learning goals. | - |
| **7** | The case-based scenarios, used throughout the distance learning, was helpful to develop my clinical knowledge | The case-based scenarios, used throughout the distance learning, was helpful, in my opinion, to develop the learners' clinical knowledge. |
| **8** | Overall, I was satisfied with the distance learning. |  |

Legend: This table shows the similarities and differences between the two surveys that were disseminated to capture the perceptions of the learners and instructors, respectively. Components 1, 2, and 8 were common between both surveys. Components 3 and 4 were meant to constitute two sides of the same coin. As for components 5 and 6, they were unique to the learners.

The second section entails the following two dichotomous questions (Yes/ No), each followed by a separate open-ended question requiring the participant to elaborate:

- The transition to the online environment, in response to the COVID-19, significantly impacted my learning (or my teaching) in these courses.
- The transition to the online environment, in response to the COVID-19, significantly impacted the courses’ structure and delivery.

As for the last section of the survey, it was meant to be exploratory to solicit for qualitative data using the following open-ended questions:

- What were some of the advantages of transitioning to distance learning?
- What were some of the challenges that you faced due to transitioning to distance learning?
- Please reflect upon aspects of the alternative modes of instruction deployed that were particularly supportive of your learning (for the learners’ distance learning) during the COVID-19 pandemic.
- What aspects of those alternative modes of instruction would you like to sustain on the long run (even after returning to regular face-to-face sessions)?

Participation in this data collection initiative was completely voluntary. The privacy and the data confidentiality of the learners were protected, and no personal identifiers were recorded. The survey was assembled throughout May 2020. In the respective academic year, the HBMCDM faculty was composed of 21 instructors and was serving a total of 63 learners.

## Data analyses

### Quantitative descriptive analyses

The quantitative data was descriptively analyzed using SPSS for Windows version 25.0. For each of the 8 quantitative components, the mean and standard deviation were calculated. An overall score of satisfaction was calculated for both stakeholders together (i.e., across the 6 components that are common to both stakeholders), along with a score of satisfaction for the learners (i.e., across all 8 components) and another one for the instructors (i.e., across the 6 components that constituted the instructors’ tool).

Since the scale used for capturing the perception of the learners and instructors was tailor-made for the purpose of this study, the validity tests of Cronbach’s Alpha and the Principal Component Analysis (PCA) were performed to ensure the internal consistency and check external variance, respectively, of the adapted tool.

For the inferential analyses, to select the appropriate tests, a test of normality was conducted for each of the 8 components, and for all three scores of satisfaction (overall, and learners and instructors). The data of each of the eight components, independently, and the overall and learners’ scores of satisfaction all turned out to be not normally distributed. As for the instructors’ data, it turned out to be normally distributed.

### Quantitative inferential analyses

Accordingly, Mann-Whitney tests were used to compare the overall score of satisfaction, and each component independently, between both groups of stakeholders (learners and instructors), and the overall score of satisfaction and learners’ score of satisfaction, between those who answered ‘Yes’ (versus those who answered ‘No’) to each of the two dichotomous questions of the second section of the survey. As for the instructors’ satisfaction score, Independent T-test was used to uncover whether, or not, this satisfaction score significantly differs between the two options of each of the dichotomous questions.

In addition, Chi-squared was used to assess any potential associations between the two dichotomous variables of the second section of the survey and the two groups of stakeholders.

Finally, the Kruskal- Wallis test was conducted to assess the extent to which the overall and learners’ scores of satisfaction can be explained by changes in the stakeholders’ perception of the components of the scores, respectively. In order to investigate the same associations for the instructors’ score of satisfaction, ANOVA was deployed.

### Qualitative analyses

The qualitative data analysis started after the conclusion of the data collection phase. The data was analyzed using Thematic Analysis by three researchers (MAH, FAR, and FO). The subjectivity of the researchers was recognized, right from the start of the analysis, to avoid affecting the integrity of the qualitative analysis trajectory. Prominent patterns were identified after thorough examination of datasets. The process was inductive, based on the constructivist epistemology. It is worth noting that the consistency, in relation to the underlying theoretical assumptions, was assured throughout the study. This iterative, interpretative approach enabled the researchers to gain a detailed understanding of the phenomenon under investigation (i.e., rapid transition to distance learning at HBMCDM at MBRU).

The process of analysis followed the six-step framework initially introduced by Braun and Clarke (2006) (14). This multi-staged approach to Thematic Analysis has been encouraged in research concerning health professions education (15). NVivo software version 12 plus (QSR International Pty Ltd, Vic, Australia) was used to code the data, and in turn expedite the categorization of the relevant text fragments.

The analysis process started with the researchers acquainting themselves with the data. The data was segmented into meaningful statements. The data collected from each of the two groups of stakeholders was handled separately. Then, as a second step, the text fragments that refer to the same aspect of the distance learning experience were compiled together, labelling each with an all-encapsulating title. Accordingly, the qualitative data was examined line-by-line, while assigning codes to text fragments, until data saturation was attained.

The researchers reflected upon areas of harmony and discord mentioned by the participants. The resulting categorization schemes of the two groups of stakeholders were mapped onto each other to compare perceptions. The same themes (more or less) surfaced in the separate analyses.

Following that, the discrete concepts, from both datasets, underwent several rounds of reflections, where the various ways by which the concepts could relate to one another were identified. This led to the generation of categories that comprehensively cover all that surfaced in relation to the two research questions, which set the stage for the researchers to work on step three. The researchers examined the categories, again, to find the best way to merge them into higher order themes.

The generated themes and categories were then reviewed as part of stage four to ensure that the data within each grouping are sufficiently similar, and data in between the clusters are distinct enough to deserve segregation. All the themes and categories were then labelled and defined to complete stage five. This constituted the basis of the study’s conceptual framework which guided the last step of reporting upon the findings.

## Results

### Quantitative analyses

#### Descriptive

Out of those 63 learners, 53 responded (i.e., response rate= 84%). As for the instructors, a total of 18 faculty members responded (i.e., response rate= 86%). Each of the 71 participants were given a unique identification number. The unique identification numbers were complimented with ‘R’ for the 53 learners, and ‘I’ for the 18 instructors (i.e., participants 1 through 53 are followed by ‘R’, and 54 through 71 by ‘I’).

The reliability score of Cronbach’s Alpha for the evaluation instrument, that captured the perception of the stakeholders was 93.3%. The percentage of the total average of the learners, instructors, and both groups of stakeholders were 82.55%, 91.13%, and 84.63%, respectively, as per Table 2.

**Table 2.**
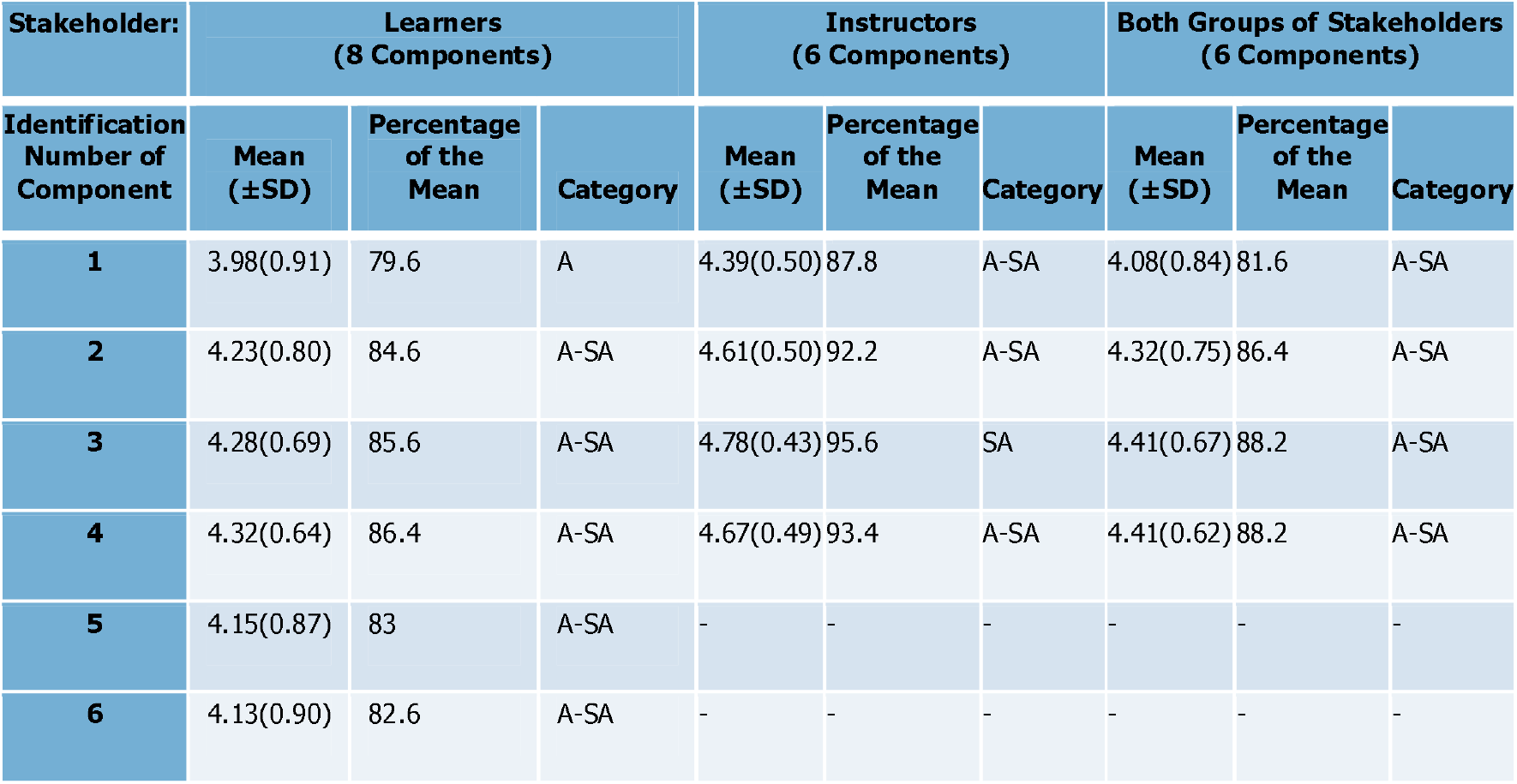
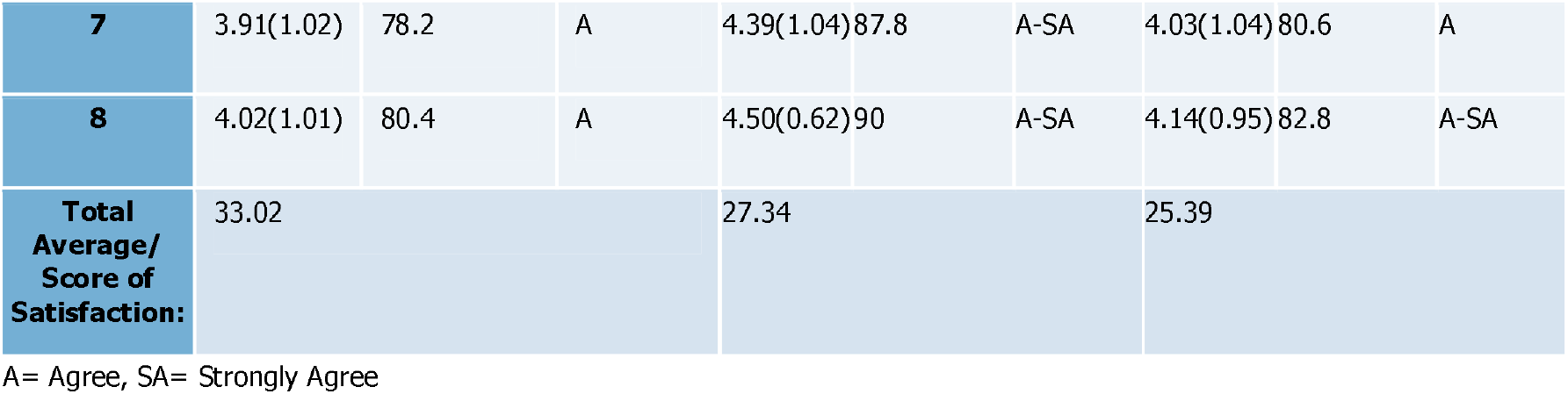
Output of descriptive quantitative analysis

According to the PCA, 90.7% of the variance can be explained by the instrument which means the instrument is not only reliable but also valid to measure what it is intended to measure.

#### Inferential

As illustrated in Figure 1, the instructors, with a mean of satisfaction of 31.84(±2.85), rated the distance learning experience higher than the learners, with a mean of satisfaction of 28.75(±5.05) (P=0.023).

**Figure 1.**
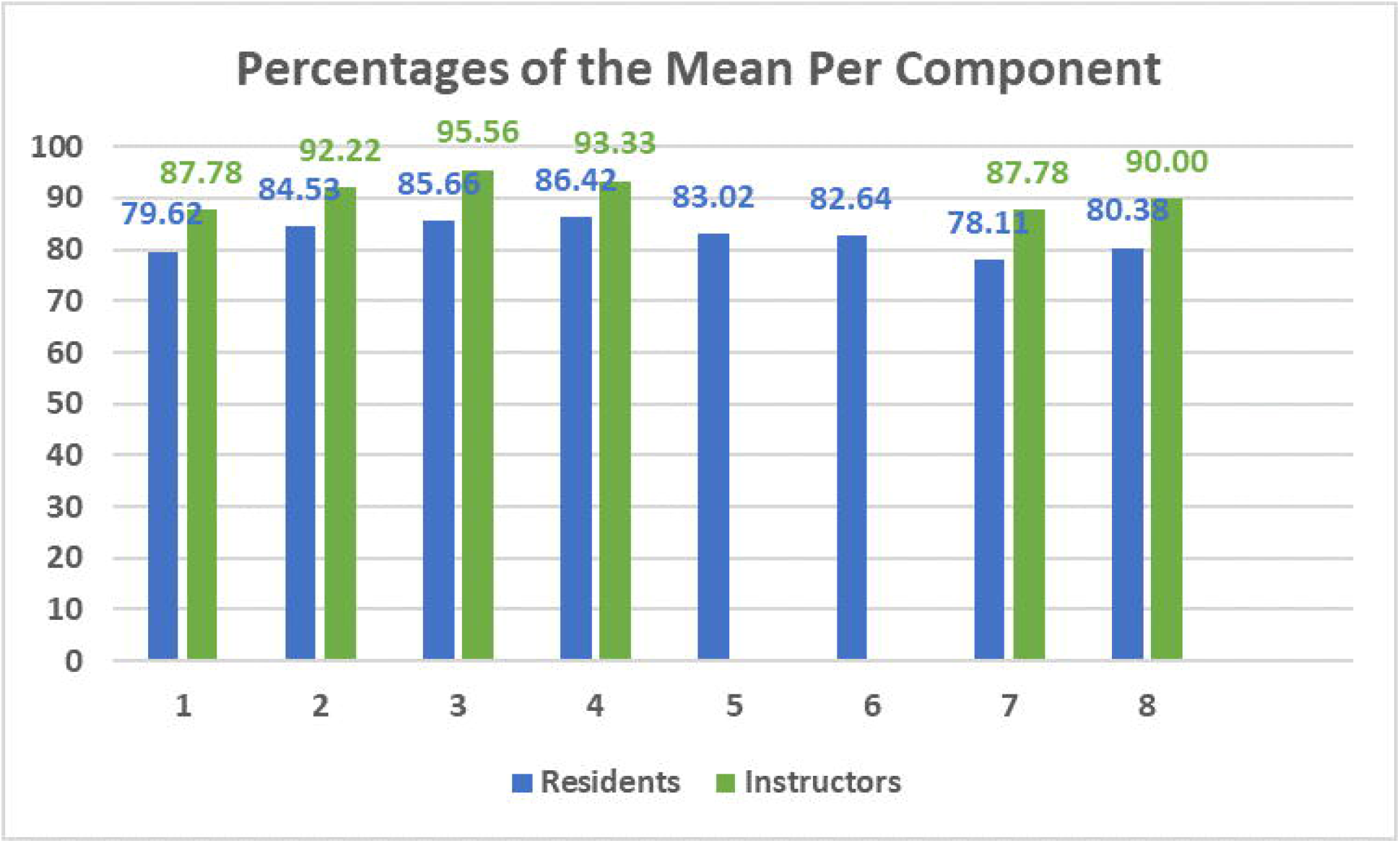
Comparison between percentages of the mean per component between learners and instructors.

The overall and learners’ scores of satisfaction were associated with all 6 components (P<0.05). As for the instructors’ score of satisfaction, it was only significantly associated with the perception of instructors regarding the following component: “the case-based scenarios, used throughout the distance learning, was helpful, in my opinion, to develop the learners’ clinical knowledge” (P=0.001).

The learners who perceived the transition not to impact the courses’ structure and delivery were significantly more satisfied (P=0.008). However, whether, or not, the stakeholders perceived the transition to impact the learning and teaching was not significantly associated with their level of satisfaction.

### Qualitative Data

The Thematic Analysis resulted in four interrelated themes: ‘Advantages’ and ‘Challenges’, ‘Modifications in Learning or Teaching’, and ‘Lessons learned and Suggestions for the Future’, as illustrated in this study’s conceptual framework (Figure 2). Within the Advantages theme, five categories surfaced: Efficiency, Convenience, Work-life Balance, Autonomy, and Cooperation. As for the Challenges theme, it encapsulated four other categories labelled as: Scope of interactions and learners’ engagement, Clinical teaching, IT limitations, and Diffusion of boundaries. The third theme: Modifications in Learning or Teaching, encapsulated four categories: Self-directed learning, Collaborative learning, Flipped Teaching, and Shortening of lectures. As for the Lessons learned and Suggestions for the Future theme, it included text fragments that refer to aspects that will be sustained, those that will be (further) leveraged, and those that require improvements.

**Figure 2.**
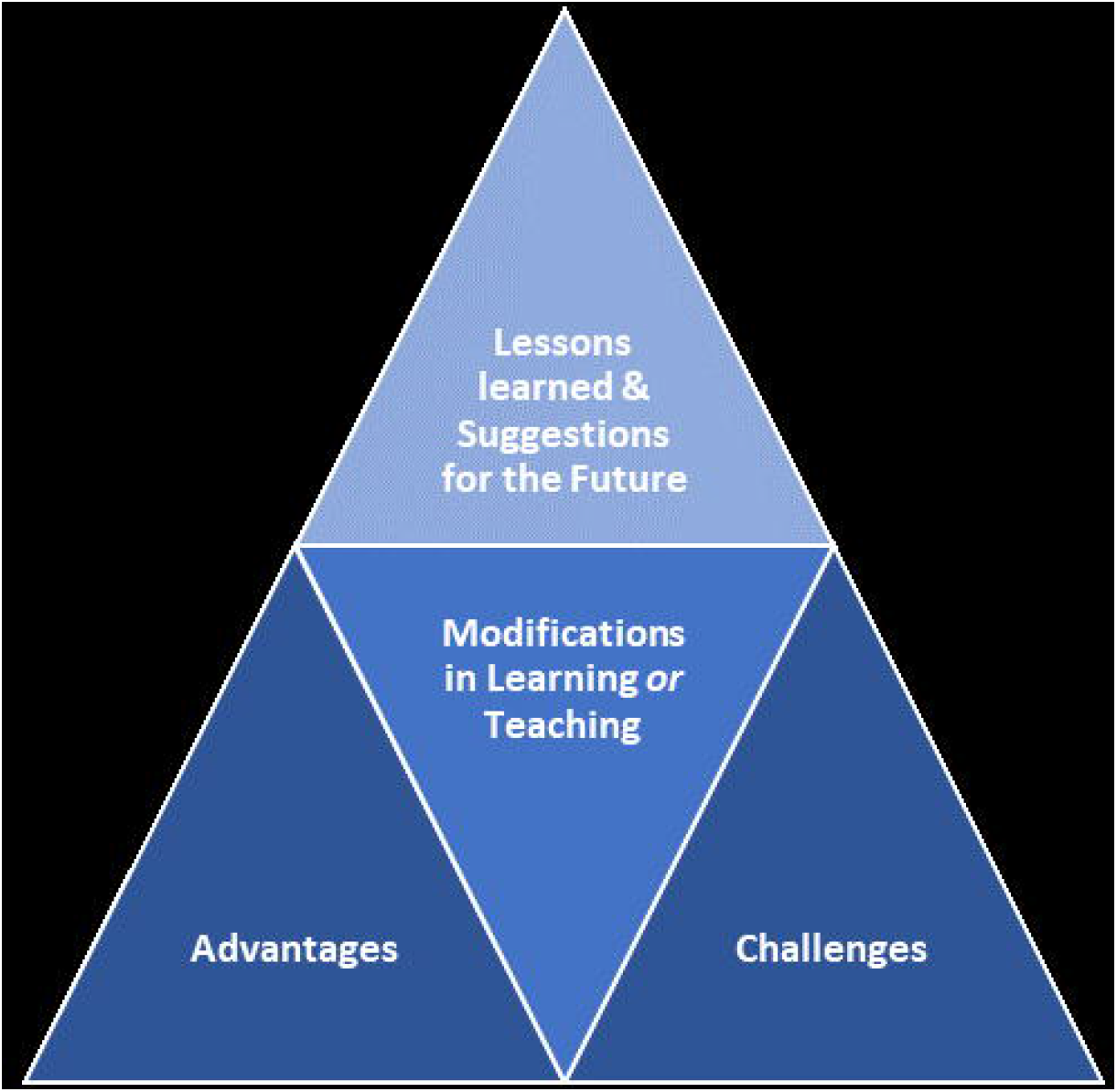
Study’s Conceptual Framework (illustrating the themes that emerged from the qualitative analyses)

### Theme 1: Advantages

This theme refers to the strengths of the rapid transition to distance learning, as perceived by the two groups of stakeholders.

### Efficiency

Both groups of stakeholders agreed that the changes accompanying the transition saved time and energy, and made the processes around the learning and teaching more efficient:

R9: “…we were getting plenty of learning materials and resources, and given enough time to ask and express our opinions, across differing classes and cases…”

F9: “…it saved time and allowed for more focus on explaining the context of the respective topics. I find the changes accompanying the rapid transition to distance learning to be very efficient…”

### Work-life balance

The stakeholders also agreed that these changes led to better work-life balance, which was particularly evident among working mothers.

R15: “…in my opinion, it has been advantageous to mothers, mostly… I have kids- I needed to support in their distance learning- they had classes and homework every day. They also needed a lot of support and encouragement to adapt to the new situation. It would not have worked out for us, as a family, if I were not at home, all the time…”

F4: “…the comfort of staying at one’s home- no commuting, remaining close to family, and the ease of connecting with any one at almost any time…”

### Convenience

The changes led to arrangements and configurations that were favorably perceived by the faculty members.

F2: “…the platform that constituted the core of the distance learning (i.e., Microsoft Teams) was even more handy and user friendly than the Learning Management System (LMS). The comments and discussions typed in the chat boxes got saved which in of itself had been a great advantage in terms of going back and attending to the topics that needed to be further discussed…”

F6: “…the ease of communication whenever needed… the ease of scheduling for and attending and participating in meetings on Microsoft Teams… there had been more flexibility and opportunities for networking, and better accessibility. Despite the external stressors due to COVID-19, the transition to distance learning made the teaching duties less stressful and more comfortable…”

### Autonomy

The learners also felt that the new normal gave them more autonomy where they had more control over their schedules and in turn managed their times better.

R2: “…it has been much more efficient than face-to-face learning, especially for people living outside Dubai, in other Emirates… by not commuting, I was able to save a lot of time and energy, which I usually spend commuting back and forth to university, and while stuck in traffic. I also made use of the time in between lectures. I got into the habit of reviewing the lectures immediately after the respective sessions, so basically the transition gave me more autonomy in managing my own schedule and making the best use of my time…”

### Cooperation

The transition to distance learning, and the virtual environment, facilitated work across physical barriers (interpersonal, and across disciplines and even nations), which was especially noticed by faculty members. This led to enhanced teamwork and better collaborations.

F2: “…the distance learning created a lot of networking and collaborating opportunities- interdisciplinary and international teamwork and teaching became much easier…”

F3: “…we all had no option but to adapt to change. The distance learning option, with all that was needed in terms of IT set-up, has been available for a long period of time, but there was resistance. Now, it is accepted and appreciated, and even preferred in some instances, among the dental education communities and institutions, across the world… this openness to change and to alternatives, and the associated flexibility are enabling plenty of collaboration and co-creating opportunities across barriers…”

### Theme 2: Challenges

This theme refers to the weaknesses and difficulties of the rapid transition to distance learning, as perceived by the two groups of stakeholders, along with struggles that they faced along this virtual journey.

### Limited scope of interactions and levels of learners’ engagement

The transition expectedly led to lessening of the scope of interactions, where people became physically, and in some instances socially, distant. In terms of knowledge exchange, this distancing required the deployment of a new set of skills, which not everyone had at the time of transition.

R6: “…the absence of the lecturer’s physical presence sometimes makes receiving and digesting the information more difficult… it is not that easy to concentrate and remain focused without seeing and interacting with the instructor in person…”

The physical distancing, integral to the changes, affected people in differing ways. The people personalities played a role. Some learners, for example, became quieter than usual and less engaged in their own learning process.

F5: “…getting all the learner to contribute especially the shy or quiet ones had been a challenge, especially that you do not see their facial expressions and nonverbals…”

### Clinical teaching

The hands-on, experiential education, which lies at the core of health professions’ education, had to be put on hold due to COVID-19 pandemic. A lot of the practical content that is usually delivered by clinical exposure got replaced by alternatives that seem to have added value but did not offer an equivalent to what the learners had missed out on.

F3: “…actually, there is no alternative to hands-on and practical activities. IT might offer solutions that effectively replace part of the experience, but that is it… health professions’ education requires clinical experience and experiential learning…”

F10: “…clinical courses, though, cannot be delivered through distance learning- this constitutes the bulk of any postgraduate dental training. Although technology gave exciting solutions to the pandemic situation, one will not become a doctor ‘remotely’…”

### Information Technology glitches and limitations

The stakeholders referred to technical glitches that occurred sporadically. They also referred to certain teaching techniques that they could not deploy online, via the Microsoft Teams platform.

F2: “…IT glitches, although rare, caused interruptions in the flow of teaching, and that usually happened when listening to learners and fitting their contributions during sessions in the grand scheme of things…”

F4: “…limitations around illustrating ad-hoc ideas to learners. Inability to draw and offer pictorial demonstrations and sketches of certain visual concepts in dentistry…”

### Diffusion of boundaries

Although, as mentioned before, distance learning and working from afar enabled crossing of physical barriers, in many instances, it led to diffusion of boundaries.

Life during the pandemic felt more like a continuum, with no clear boundaries, for the learners and faculty members.

R4: “… finding a quiet place at home was not easy… distraction when family members, especially the kids, pass-by and require my attention assuming I am not engaged or busy with something else… at the end of the day, they are kids- it is difficult for them to realize I am doing work but from home…”

R9: “…distractions at home, especially that all other members of the family were also working and/ or studying from home. Having kids around is difficult…”

### Theme 3: Modifications in Learning or Teaching

This theme refers to how the two groups of stakeholders modified their approaches to learning or teaching in order to adapt to the rapid transition to distance learning. The novel learning and teaching environment became conducive to the favorable behavioral and attitudinal changes, where both groups of stakeholders seem to have organically grown and developed, personally and professionally, from this experience. The perceptions of the two groups map onto each other, like two sides of the same coin.

### Self-directed learning

The learners exhibited more proactiveness throughout the distance learning. learners took the initiative to and went out of their way to foster their learning throughout the virtual experience. They attributed the tendency of self-directed learning to having more space and time at hand.

R14: “…I attended free dental webinars, from all over the world- that was amazing…”

R8: “…the space that this transition led to enabled us to resort to and use more resources than usual to study… access to journals and articles helped me a lot in better informing myself in regards to what we were learning as part of the program. The resources have always been there- but because we were distant, and had more space and energy at hand, we used them to our advantage…”

### Collaborative learning (vis-à-vis group-based learning)

Teaching became more group-based and learners were more collaborative. The instructors assigned more group-based exercises, and the learners were more open to sharing resources and engaging with each other; efforts to come together and co- create were evident on both sides.

R17: “…transition to team-based learning had been conducive to our learning, in my opinion. Working in groups had been better because we divided the workload and assigned responsibilities. Each one worked independently, across differing roles, towards a common goal. We became more productive, as a team…”

F7: “…learners, from differing cohorts and programs, used to have difficulties in setting mutually-convenient meeting times to work together (due to their different timetables). This experience opened doors for new collaboration opportunities…”

### Flipped Teaching

The faculty members resorted to flipped teaching, more often, where the direct instruction moves from the group to the individual learning space. This group space is transformed into an interactive learning environment where the instructor guides the learners as they apply concepts and engage creatively in the subject matter.

F1: “…we adapted flipped learning models, which I think is better than the traditional ‘spoon feeding’ learning, especially for postgraduate teaching which our learners were used to.”

### Shortening of lectures

Along the same line, most of the sessions got shortened.

R1: “…the longer the session, the less we are able to remain focused. We became way more attentive during the lectures that were reduced in duration…”

F7: “…almost all lectures were shortened, since two-hour distance learning lectures did not work for us…”

### Theme 4: Lessons learned & Suggestions for the Future

This theme encapsulated what the stakeholders acquired from and their recommendations due to this first-hand experience with distance learning. The institutional knowledge generated from this experience better prepared the stakeholders for upcoming rounds of distance learning. Due to this experience and what the two groups of stakeholders gained from it, the upcoming round of distance learning is expected to improve.

### Aspects that will be sustained

Some of the aspects worked well and will be maintained as is.

R7: “…distance learning is beneficial for learners who cannot attend lectures, in person, on campus. Instead of missing classes, they can attend them online…”

R15: “…teleconsultations via Microsoft Teams (that we started using, with the patients, only after COVID-19) offered amazing learning experiences, and is going to stay with us in the Orthodontics department, irrespective of how the modes of delivery shape-up…”

R13: “…the additional case-presentations exposed us to new cases that we do not typically see in clinical training. Getting more frequently exposed to CBD enabled us to examine certain cases more and elaborate upon them…”

### Aspects that will be (further) leveraged

Other aspects worked well and were discovered to be worth capitalizing upon in a systematic manner. These aspects exhibited potential that is worth exploiting to maximize the learning and teaching experiences.

R9: “…we recommend for us to have more joint sessions with postgraduate learners in other universities, to be informed about and in turn to attend more webinars offered by other universities, to invite more learners from other universities to attend webinars offered by our university, and to conduct more interdisciplinary discussion sessions within the university…”

F2: “…engaging more with learners and faculty members in other institutions, across the world…”

### Aspects that require improvements

There were also aspects that were identified to require improvements. In other words, matters did not work well and/ or entailed noticeable opportunity for improvement.

R12: “…having access to recordings of the live lectures, with the PowerPoint together (Picture-in-Picture), would be helpful… we can go back and listen to the instructor’s comments at any time…”

R6: “…to use more of pre-existing educational videos such as procedural videos or clinical scenarios during the lectures… it makes the information more digestible…”

## Discussion

This study showed that both groups of stakeholders: learners and instructors, were quite satisfied with the rapid transition to distance learning due to COVID-19. The transition was characterized by several advantages as perceived by the stakeholders. The continued learning and teaching via the online platforms saved time and energy, especially around commuting back and forth between one’s home and campus, and increased the efficiency of the associated processes, all of which were considered more convenient for the stakeholders relative to the face-to-face configuration. This led to enhancing the work-life balance for the learners and the instructors. The learners favored having more control over their schedules, and both parties were happy with the increased cooperation across the board. This is in concordance with previous literature that highlighted enabling self-paced learning, and allowing for more time and space flexibility as prominent advantages of distance learning (16, 17). Whereas another study, demonstrated an undesirable result of increased autonomy among learners who are not self-regulated and/ or not equipped with time management skills (18). Accordingly, supporting learners and instructors in developing those competencies would help them in maximizing their learning and teaching experiences, while striking a better work-life balance.

Although both parties scored highly in terms of satisfaction with the distance learning, the group of instructors turned out to be significantly more satisfied than the group of learners. It was previously suggested in the literature among the factors that influence the level of satisfaction of online teaching for instructors are self-gratification, intellectual challenge, interest in using technology, and the associated professional development opportunities (19). The instructors’ overall level of satisfaction was associated with their satisfaction with the additional CBD sessions. The same component variable was also an antecedent to the overall score of satisfaction of learners. The qualitative data analysis uncovered the same findings, where both groups of stakeholders would like to maintain additional online CBD sessions in the future. The overall learners’ score of satisfaction was not only associated with their perception of the case-based learning, but also with all the rest of the components of the adapted tool which proved to be valid and transferrable to other similar contexts. Moreover, learners who perceived the transition to be seamless, without impacting the courses’ structure and delivery, were significantly more satisfied, which is why it is important to work towards instilling a culture of change when planning for systematic transitioning to distance learning. It is also important to ensure providing sufficient technical support along the way. This will decrease the level of resistance which was previously proven to enhance the overall distance learning experience (20-22).

The stakeholders also pinpointed challenges that they faced in their distance learning experiences. The perceived added value of CBD did not substitute for hands-on experience. The distance learning was satisfying for non-clinical teachings only. As for the clinical training, it lagged. Resources, which offer equivalents to real-life experiences, clinical trainings, and/ or interactions with patients, were non- existent.

Another prominent challenge was the noticeable decrease of in-class contributions; the learners highlighted how their concentration span seemed to become shorter after the transition, and the instructors reflected upon the difficulties that they faced in keeping the learners focused and engaged. Along the same lines, it is established in the literature that maintaining the level of interaction and keeping learners engaged are among the most prominent challenges instructors face in online teaching (22, 23). This challenge was further exacerbated whenever the stakeholders faced technical glitches. Fortunately, those incidences were rare, as reported by several stakeholders, but it is important to keep this potentiality in mind because at the end of the day the IT platform constitutes the medium through which the whole experience occurs. Any hurdles, on that front, would result in interruptions to the online learning and teaching experience.

The learners and instructors noticed that (and in turn reported upon how) they organically modified their learning or teaching styles, respectively, to adapt to the circumstances and to maximize the experiences. The external consequences led to behavioral changes, for both parties. In the literature, it is established that choices of pedagogical approaches by instructors are expected to affect learners’ learning approaches (24). Some instructors reported shortening of their lectures. Others mentioned resorting to flipped learning approaches which entailed making use of Questions and Answers, discussions, quizzes (i.e., formative assessments to ensure timely attainment of the intended learning outcomes of the sessions), and providing of feedback throughout the online sessions. The learners, in turn, deployed more self-directed learning, which is desired in fostering life-long learning. Collaborative learning increased, as well; more group activities were assigned by the instructors, and the learners became more likely to resort to supporting each other, sharing of resources, and working together towards common goals. All of which are in alignment with the principles of connectivism (25). This can be further emphasized by consideration of virtual-based situated learning, where content in the online medium needs to be engaging, the environment needs to be contextualized, cultivation of participation and fostering active learning become the priorities, and forming and nurturing community-of-practice is recommended (26).

A lot of insights and lessons learned were acquired from the rapid transition to distance learning which can be effectively assimilated and reinforced with pre-existing evidence to better integrate and sustain distance learning. Blended learning has been implemented, with favorable outcomes, in other post-graduate dental schools (27-29). Considering the high acceptance of distance education among this study’s learners and instructors, effective integration of blended education, complemented with flipped classroom in certain subjects, holds the potential of substantial benefits for both groups of stakeholders. The concept of blended education can incorporate asynchronous and synchronous learning elements (30) (31). As a result of the pandemic, teleconsultation was employed, in some of the programs, and was appreciated for offering the learners opportunities to acquire new methods of consultation. In relation to the clinical exposure, the massive appreciation around the CBD, and the debriefing sessions after the presentations of clinical scenarios, can be leveraged to enhance critical thinking, decision making, and clinical reasoning skills (32, 33). In addition, this andragogy, that proved to be of substantial added value to the online learning and teaching in the college under investigation, can be systematically integrated into the curriculum for it to evolve into full-fledged case-based learning.

Besides all the challenges that it brought along; the COVID-19 pandemic created plenty of novel opportunities in the worldwide dental community. To make up for the distance and to make good use of the space that this period has created, the dental educators became more engaged via virtual webinars and online conferences. Those opportunities were open and easily accessible by all members of the community-at-large, where knowledge and resources sharing among the stakeholders expanded in scale and scope. A network of dental education institutions was formed, which nurtured international collaborations. Bridging between educational institutions and clinical practitioners was also evident. This interconnected learning community is expected to support post-graduate learners in their future job search and academic collaborations. It also encourages the learners and instructors to come-up with novel, innovative pedagogical techniques through assimilating materials that are readily available online (e.g., case-scenario banks or educational procedural videos), based upon the constructivist theoretical underpinnings (via strategies such as: inculcation and integration). Along the same lines, among the wide array of resources that learners get access to, are videos of procedures which enable clinicians to practice the associated set of skills prior implementing it for a patient, all of which are desired in any curriculum but seldom occurs(34, 35).

This study is characterized by a few limitations. In alignment with the principles of the Institutional Research function, complete anonymity of the participants was maintained. Therefore, the gender, age, and the current level in the respective programs of the participants were not recorded. It would have been interesting to know if the satisfaction and the perceived impact of the transition are associated with those demographic variables. Also, the qualitative data offered a lot of insights that could have been further explored with alternative data collection tools (e.g., focus group sessions). Moreover, although the focused study design enabled the development of thorough insights, the generalizability of the findings is limited to institutions that are contextually and characteristically like MBRU. This limitation is further pronounced given the exceptionality of the times of the COVID-19 pandemic (on all fronts). It goes without saying that both groups of stakeholders faced major changes in their personal and social lives. Their day-to-day life was majorly disrupted, and they were all under a lot of pressure due to the halting of the clinical training which, in principle, entails 60% of the postgraduates’ time. This left the stakeholders with more time to study and to engage in other virtual educational activities which might not remain the case after resuming the clinical learning. Therefore, any decision on the changes of the curriculum towards blended learning needs to be tailored in accordance with the reality of post-pandemic circumstances. Finally, this study evaluated the official platforms of learning. It was evident, though, that unofficial learning tools such as Social Media Applications (SMA) were of added value to the learning and teaching. Properly understanding how such learning tools can be maximized is important when planning for distance learning.

It would be great to build upon this study in the following directions. To start with, it is worth exploring the long-term effect of this unprecedented abrupt change in educational method, due to COVID-19 pandemic, on the learners as they progress. Moreover, it would be useful to develop a contextualized competency-based model that would constitute the foundation for instructors’ professional development. It would be interesting for the college to adapt action research to develop, with thorough engagement of the stakeholders, contingency learning, and teaching plans for such times of crisis. Finally, the distance learning started after the spring break, which means the first-year learners had about 6 months of face-to-face learning, which enabled them to build rapport with their instructors and colleagues. It would be interesting to investigate if first-year learners, who sign-up for blended learning from the beginning of the academic year, would perceive matters differently.

## Conclusion

The abrupt transition to distance learning, due to COVID-19, was perceived favorably by the involved stakeholders at the respective college. This unexpected change entailed overcoming plenty of challenges, but also uncovered substantial opportunities that are worth capitalizing upon in health professionals learning and teaching. The lessons learned, and the first-hand knowledge that stakeholders acquired from the reaction to the onset of the pandemic, can be leveraged to innovatively develop and reinforce post-graduate dental curriculums. This reality-check has put stakeholders of higher education in a position to work together and share knowledge. This in turn will enable them to be better prepared for any such transitions and will equip the learners and educators with the skills to maximize the learning experiences and ensure educational continuity.

## Data Availability

Data can be provided upon request.

## Supporting information captions

Appendix 1: Distance learning survey for learners and instructors.

Appendix 2: Surveys’ raw data

